# Cardiac Rehabilitation is Associated with Improved Clinical Outcomes in Patients with Chronic Total Occlusions: A Large-Scale, Propensity-Matched Analysis

**DOI:** 10.64898/2026.03.25.26349342

**Authors:** Chirayu R. Shukla, Charles D. Miks, Pranav Puri, Grant K. Ozaki, Alex Cuskey, Hunter Frederiksen, Joseph Phillips, Philip Horwitz, Paari Dominic, Vikram Sharma

**Author notes:** **Ethical Approval:** The University of Iowa Health Care IRB Chair or Chair Designee has determined that this project does not meet the regulatory definition of human subject research and does not require review by the IRB, because this activity is based on evaluation of de-identified outcome data using TriNetX. **Corresponding Author:** Chirayu Shukla BS, 2869 Spring Rose Circle, Coralville, IA 52241.

## Abstract

**Background:** Chronic total occlusions (CTOs) are a common manifestation of coronary artery disease (CAD) and are associated with increased long-term mortality. While successful CTO revascularization improves symptoms and quality of life, a consistent mortality benefit has not been demonstrated in randomized trials. Outpatient cardiac rehabilitation (CR) has proven benefits in improving functional status, exercise capacity, and quality of life in patients with CAD, yet its impact on CTO patients has not been well studied.

**Objective:** To evaluate the association between CR and long-term outcomes in CTO patients.

**Methods:** Using the TriNetX Research Network, we analyzed de-identified patient data from 75 healthcare organizations using ICD codes. The study population included patients with CTO who started CR within 3 months of diagnosis vs patients with CTO who did not engage in CR. A secondary analysis was also conducted, which excluded patients with other indications for CR, including prior coronary artery bypass grafting (CABG) and prior or concurrent percutaneous coronary interventions (PCI).

**Results:** Of 167,176 CTO patients, 10,021 enrolled in CR, including 1,608 without another CR indication. Patients were propensity-matched for independent risk factors for mortality. After 5 years, CR participation was associated with a significant reduction in mortality (HR 0.68; 95% CI, 0.61–0.75; p < 0.0001). This benefit was preserved even after excluding prior revascularization (HR 0.81; 95% CI, 0.67–0.99; p < 0.036).

**Conclusion:** This study demonstrates that cardiac rehabilitation is associated with improved long-term survival in patients with CTOs.

## Introduction

Chronic total occlusion (CTO) of the coronary artery, defined as 100% occlusion of a coronary artery for at least three months, is found in at least 25 to 33% of patients undergoing a coronary angiography¹. Patients with CTOs tend to have greater symptom burden, impaired quality of life, and worse long-term prognosis compared with those who have non-occlusive coronary disease.

Although there have been advances in percutaneous coronary intervention (PCI) and surgical techniques, the long-term benefits of CTO revascularization remain debated^1^. While some observational studies have suggested improved survival among patients with successful revascularization ^2^, randomized trials have failed to show reductions in major adverse cardiac events (MACE)^3^. Recent long-term studies have also yielded conflicting results, with some suggesting benefit from revascularization for concomitant CTOs after a myocardial infarction^4^, while others have demonstrated no difference between CTO and non-CTO revascularization groups^5^. Although there is evidence that successful PCI for CTO can improve exercise capacity and quality of life,1,11,12, durable effects on hard outcomes, such as all-cause mortality, remain unclear.

In addition, CTO PCI carries substantial procedural complexity and a higher risk of complications, including coronary perforation, cardiac tamponade, radiation exposure, and contrast-induced nephropathy (REF). Although these are reduced in high-volume, specialized centers, they remain important considerations during patient selection. Several clinical and angiographic characteristics have been identified as negative predictors of procedural success. Clinically, prior myocardial infarction, systolic heart failure, peripheral vascular disease, hypertension, diabetes, and chronic kidney disease portend worse outcomes. Angiographic characteristics of multivessel disease, severe calcification, proximal cap ambiguity, long occlusion length, and significant vessel tortuosity (REF) similarly predict worsened outcomes. Unfortunately, many patients in the CTO population possess multiple risk factors that preclude CTO intervention. Even in patients who undergo successful CTO intervention, some will have refractory symptoms. For this reason, alternative modalities to improve QOL and mortality in the CTO population are needed.

Exercise-based cardiac rehabilitation (CR) is a well-established intervention that combines supervised exercise training, risk factor modification, education, and behavioral counseling. Systematic reviews and meta-analyses have shown that CR reduces cardiovascular mortality and hospital readmissions, while also improving exercise capacity and quality of life ^6,7^ in traditional CAD patients. Furthermore, long-term follow-up suggests that CR after PCI has benefits that persist for more than a decade after revascularization, with lower rates of MACE and improved survival ^8^. Reflecting this robust evidence base, current guidelines emphasize CR as a core component of secondary prevention programs in those with CAD^9^⁻^10^.

Despite the established benefits, CR remains underutilized, with only 19–34% of eligible patients participating in programs^13^. There is also a lack of evidence that specifically examines the long-term outcomes or benefits of CR in patients with CTOs. Given the high symptom burden and uncertainty of intervention in patients with CTO, along with the well-documented advantages of CR in other coronary populations, this represents an important gap in current literature.

The aim of this study was to evaluate long-term outcomes among patients with CTOs who did vs those who did not participate in CR, using data from the TriNetx Research Network. We hypothesized that CR would be associated with improved survival and reduced rates of MACE compared with no CR participation.

## Methods

We designed a retrospective cohort study using data from the TriNetX research network, a federated, cloud-based platform providing access to de-identified electronic health record data from participating healthcare organizations. We identified adult patients (≥18 years) with a CTO diagnosis using ICD-10 code *I25.82* or equivalent ICD-9 terminology. Participation in cardiac rehabilitation (CR) was identified using CPT (93797, 93798) and HCPCS (G0422, G0423) codes. The index event was defined as the first CTO diagnosis in all groups, and patients with any CR within 12 months prior to the index event were excluded. Patients were then stratified into two groups comparing those who started CR within 3 months after the index event and those who did not. Because CR is known to improve outcomes following coronary intervention (PCI or CABG), a secondary analysis excluding patients with prior intervention 6 months prior to 3 months after initial diagnosis was performed to ensure CR benefits extend specifically to the CTO population.

Baseline characteristics for high-risk features and demographic variables influencing outcomes and/or candidacy for CR were summarized using means (standard deviation) for continuous variables and counts (percentages) for categorical variables (Table 2). Propensity score matching was performed to reduce confounding between CR and non-CR groups. The primary outcome was all-cause mortality at 5 years following the index event. Secondary outcomes included repeat coronary revascularization (PCI or CABG) and major adverse cardiac events (MACE), defined as a composite of all-cause mortality, nonfatal myocardial infarction, and stroke, with A landmark analysis was performed to mitigate immortal time bias in the CR group, with outcomes measured starting 3 months after the index event until death, loss to follow-up, or the end of the 5-year follow-up period, whichever occurred first. Time-to-event outcomes were evaluated using Kaplan–Meier survival analysis, with differences assessed using log-rank tests. Hazard ratios with 95% confidence intervals were estimated using the Cox proportional hazards model. Patients were censored at their last known follow-up, and those with outcomes prior to the defined time window were excluded from analyses. All analyses were conducted within the TriNetX analytics platform (Figure 2).

**Figure 1.**
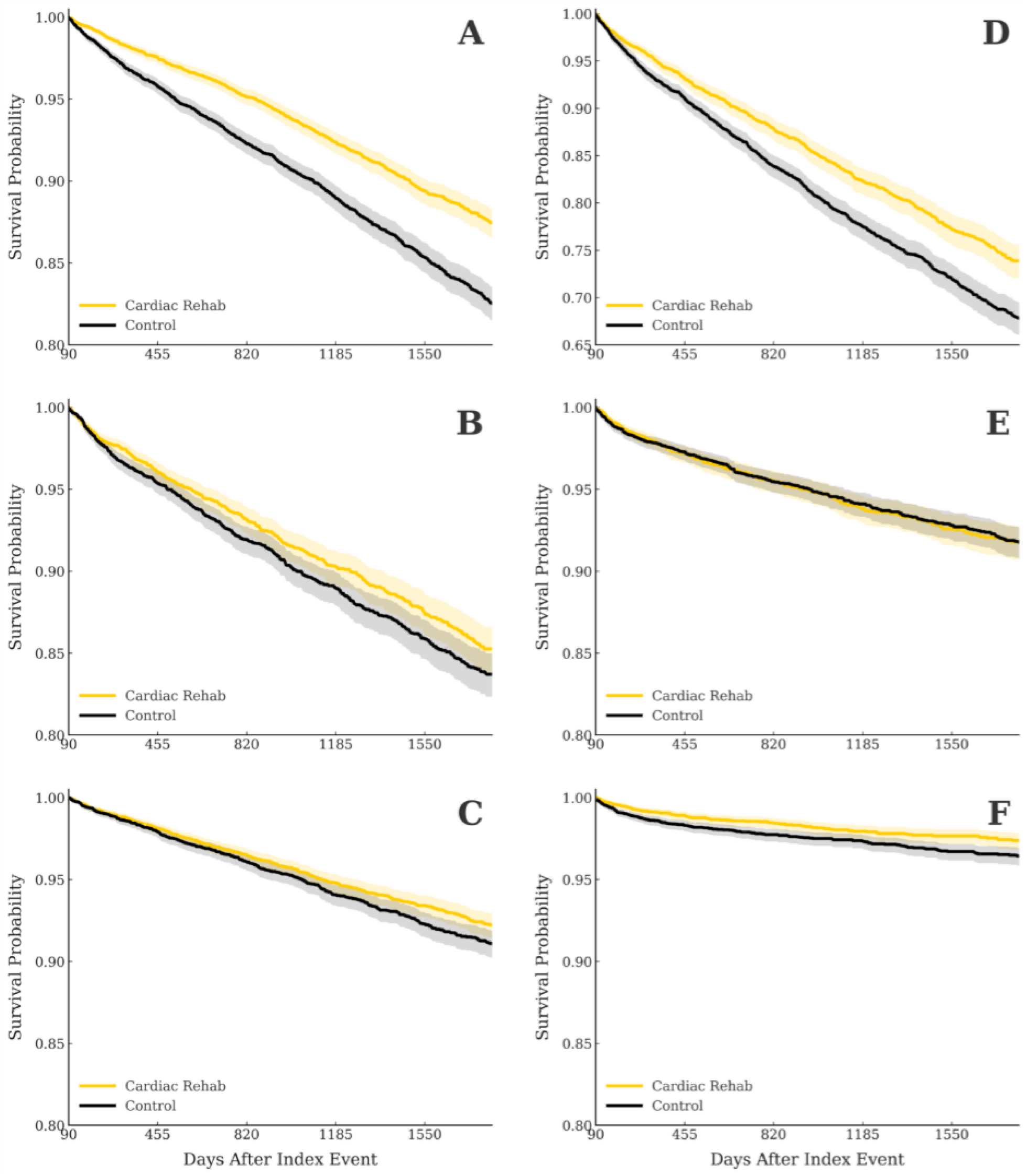
Kaplan-Meier event-free survival curves for all-cause mortality (A), acute MI (B), stroke (C), MACE (D), PCI (E), and CABG (F) from 90 days to 5 years following CTO diagnosis. Patients undergoing CR (yellow) are compared to those who did not (black). Shaded regions indicate 95% confidence intervals. The CR group (yellow) demonstrated reduced all-cause mortality (A), acute MI (B), stroke (C), MACE (D), and CABG (F). *CABG = Coronary Artery Bypass Graft; CR = Cardiac rehabilitation; CTO = Chronic Total Occlusion; MACE = Major Adverse Cardiac Events; MI = Myocardial Infarction; PCI = Percutaneous Coronary Intervention*

**Figure 2.**
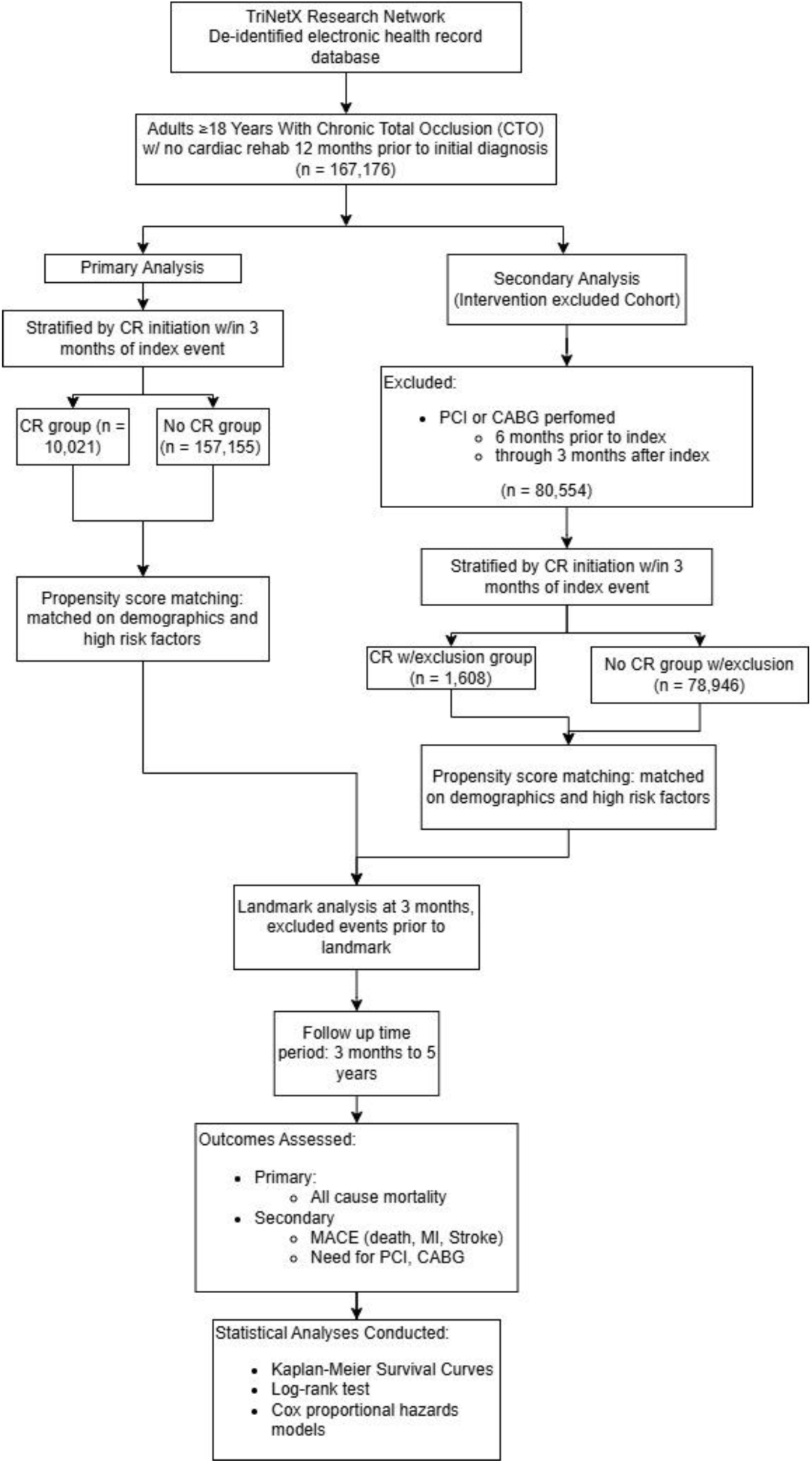
Methods flow chart for primary and secondary analyses. The primary analysis is on the right side, with the secondary analysis on the left. Prior CABG and PCI (6 months prior) through 3 months after the index event were excluded from the secondary analysis. *CABG = Coronary Artery Bypass Graft; CR = Cardiac rehabilitation; CTO = Chronic Total Occlusion; MACE = Major Adverse Cardiac Events; MI = Myocardial Infarction; PCI = Percutaneous Coronary Intervention*

**Table 1.**
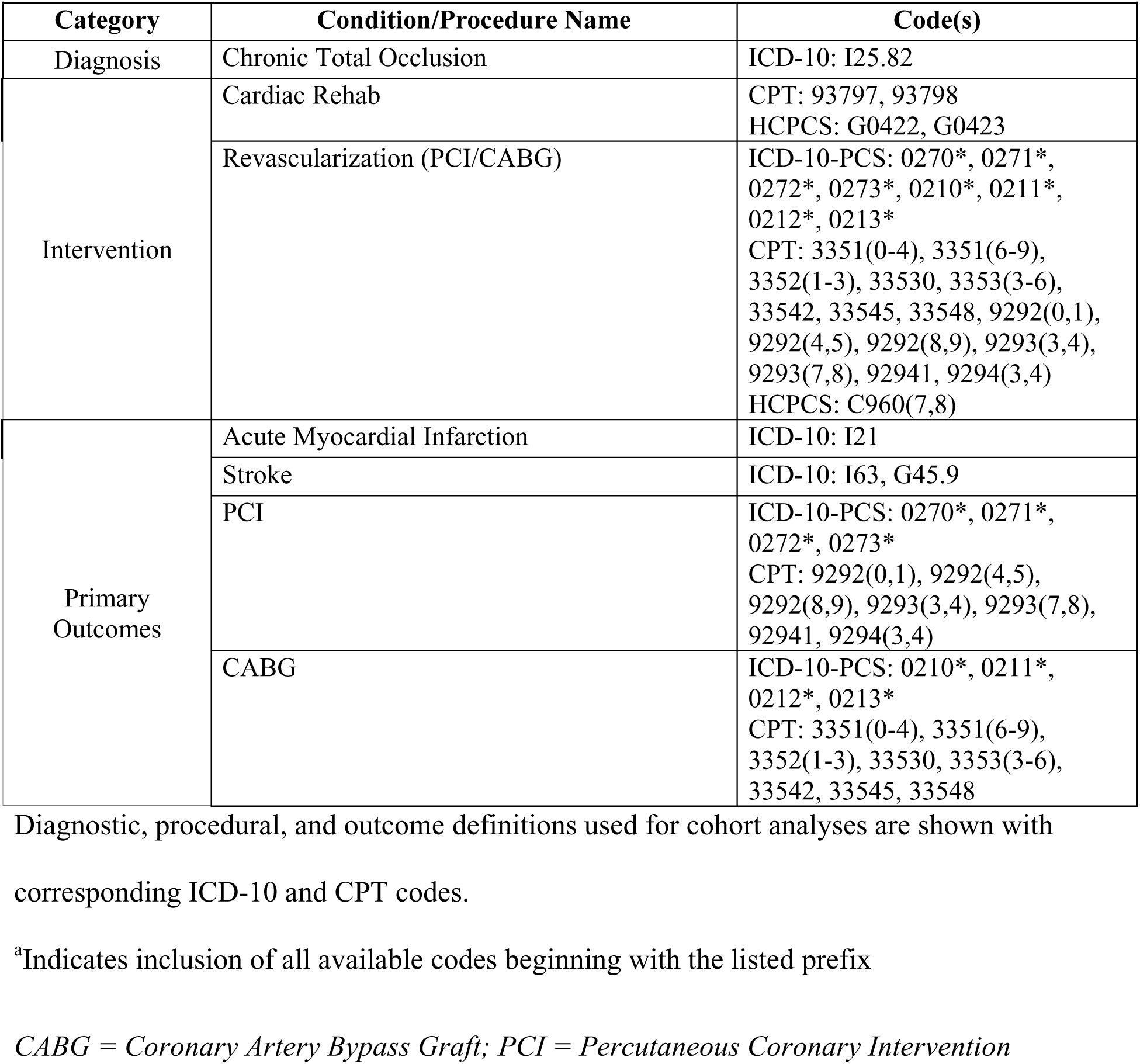
Diagnostic Codes Used for Cohort Analysis.

| Category | Condition/Procedure Name | Code(s) |
| --- | --- | --- |
| Diagnosis | Chronic Total Occlusion | ICD-10: I25.82 |
| Intervention | Cardiac Rehab | CPT: 93797, 93798<br>HCPCS: G0422, G0423 |
|  | Revascularization (PCI/CABG) | ICD-10-PCS: 0270*, 0271*, 0272*, 0273*, 0210*, 0211*, 0212*, 0213*<br>CPT: 3351(0-4), 3351(6-9), 3352(1-3), 33530, 3353(3-6), 33542, 33545, 33548, 9292(0,1), 9292(4,5), 9292(8,9), 9293(3,4), 9293(7,8), 92941, 9294(3,4)<br>HCPCS: C960(7,8) |
| Primary Outcomes | Acute Myocardial Infarction | ICD-10: I21 |
|  | Stroke | ICD-10: I63, G45.9 |
|  | PCI | ICD-10-PCS: 0270*, 0271*, 0272*, 0273*<br>CPT: 9292(0,1), 9292(4,5), 9292(8,9), 9293(3,4), 9293(7,8), 92941, 9294(3,4) |
|  | CABG | ICD-10-PCS: 0210*, 0211*, 0212*, 0213*<br>CPT: 3351(0-4), 3351(6-9), 3352(1-3), 33530, 3353(3-6), 33542, 33545, 33548 |

Diagnostic, procedural, and outcome definitions used for cohort analyses are shown with corresponding ICD-10 and CPT codes.
<sup>a</sup>Indicates inclusion of all available codes beginning with the listed prefix
*CABG = Coronary Artery Bypass Graft; PCI = Percutaneous Coronary Intervention*

**Table 2.** Baseline Characteristics of CTO Patients Starting CR.

|  |  | Before Propensity Score Matching |  |  | After Propensity Score Matching |  |  |
| --- | --- | --- | --- | --- | --- | --- | --- |
| Characteristics | Codes* | +CR | -CR | p-value | +CR | -CR | p-value |
| Number of Patients |  | 10,021 | 157,155 |  | 10,020 | 10,020 |  |
| Age: mean yrs ± SD (continuous) | | 65.5 ± 10.9 | 67.6 ± 11.2 | < 0.0001 | 65.5 ± 10.9 | 65.5 ± 11.3 | 0.9547 |
| Male |  | 7706 (76.9) | 113044 (71.9) | < 0.0001 | 7705 (76.9) | 7697 (76.8) | 0.8934 |
| White |  | 8074 (80.6) | 114695 (73.0) | < 0.0001 | 8073 (80.6) | 8098 (80.8) | 0.6546 |
| Black or African American |  | 802 (8.0) | 13185 (8.4) | 0.1742 | 802 (8.0) | 799 (8.0) | 0.9377 |
| Hispanic or Latino |  | 530 (5.3) | 9844 (6.3) | < 0.0001 | 530 (5.3) | 494 (4.9) | 0.2481 |
| Asian |  | 328 (3.3) | 8542 (5.4) | < 0.0001 | 328 (3.3) | 298 (3.0) | 0.2231 |
| Other Race |  | 216 (2.2) | 4313 (2.8) | 0.0004 | 216 (2.2) | 232 (2.3) | 0.4446 |
| American Indian or Alaska Native |  | 41 (0.4) | 635 (0.4) | 0.9387 | 41 (0.4) | 43 (0.4) | 0.8269 |
| Native Hawaiian or Other Pacific Islander |  | 25 (0.3) | 2691 (1.7) | < 0.0001 | 25 (0.3) | 43 (0.4) | 0.0288 |
| Hypertension | I10-I15 | 7102 (70.9) | 110123 (70.1) | 0.0939 | 7101 (70.9) | 7215 (72.0) | 0.0746 |
| Hyperlipidemia | E78.5 | 6025 (60.1) | 91130 (58.0) | < 0.0001 | 6024 (60.1) | 6094 (60.8) | 0.3118 |
| Diabetes mellitus | E08-E13 | 3767 (37.6) | 63274 (40.3) | < 0.0001 | 3767 (37.6) | 3840 (38.3) | 0.288 |
| Overweight and obesity | E66 | 2841 (28.4) | 39029 (24.8) | < 0.0001 | 2841 (28.4) | 2758 (27.5) | 0.1913 |
| Tobacco use | Z72.0 | 755 (7.5) | 10751 (6.8) | 0.008 | 754 (7.5) | 661 (6.6) | 0.0103 |
| Obstructive sleep apnea | G47.3 | 1608 (16.1) | 22873 (14.6) | < 0.0001 | 1607 (16.0) | 1549 (15.5) | 0.2607 |
| Systolic heart failure | I50.2 | 1229 (12.3) | 28016 (17.8) | < 0.0001 | 1229 (12.3) | 1186 (11.8) | 0.3508 |
| Diastolic heart failure | I50.3 | 755 (7.5) | 18185 (11.6) | < 0.0001 | 755 (7.5) | 740 (7.4) | 0.6867 |
| Cardiomyopathy | I42 | 950 (9.5) | 20731 (13.2) | < 0.0001 | 950 (9.5) | 918 (9.2) | 0.4369 |
| Atrial fibrillation and flutter | I48 | 1319 (13.2) | 30743 (19.6) | < 0.0001 | 1319 (13.2) | 1369 (13.7) | 0.3000 |
| Acute myocardial infarction | I21 | 2352 (23.5) | 42065 (26.8) | < 0.0001 | 2351 (23.5) | 2310 (23.1) | 0.4930 |
| Peripheral vascular disease | I73.9 | 1157 (11.6) | 25778 (16.4) | < 0.0001 | 1157 (11.6) | 1139 (11.4) | 0.6897 |
| Cerebral infarction | I63 | 656 (6.6) | 12900 (8.2) | < 0.0001 | 656 (6.6) | 630 (6.3) | 0.4536 |
| Transient cerebral ischemic attacks | G45.9 | 380 (3.8) | 6834 (4.4) | 0.0078 | 380 (3.8) | 312 (3.1) | 0.0085 |
| Chronic obstructive pulmonary disease | J44 | 976 (9.7) | 25606 (16.3) | < 0.0001 | 976 (9.7) | 964 (9.6) | 0.7744 |
| Anemia, unspecified | D64.9 | 1363 (13.6) | 29983 (19.1) | < 0.0001 | 1363 (13.6) | 1334 (13.3) | 0.5483 |
| CKD stage 3 | N18.3 | 1004 (10.0) | 20804 (13.2) | < 0.0001 | 1004 (10.0) | 968 (9.7) | 0.3932 |
| CKD stage 4 | N18.4 | 224 (2.2) | 7077 (4.5) | < 0.0001 | 224 (2.2) | 224 (2.2) | 1.0000 |
| CKD stage 5 | N18.5 | 99 (1.0) | 3696 (2.4) | < 0.0001 | 99 (1.0) | 96 (1.0) | 0.8291 |
| ESRD | N18.6 | 219 (2.2) | 8089 (5.2) | < 0.0001 | 219 (2.2) | 226 (2.3) | 0.7372 |
| Osteoarthritis | M15-M19 | 2788 (27.8) | 37668 (24.0) | < 0.0001 | 2787 (27.8) | 2803 (28.0) | 0.8010 |
| Dementia | F03 | 68 (0.7) | 2667 (1.7) | < 0.0001 | 68 (0.7) | 61 (0.6) | 0.5364 |
| Depression | F32 | 1451 (14.5) | 23412 (14.9) | 0.2524 | 1451 (14.5) | 1358 (13.6) | 0.0584 |
| Generalized anxiety disorder | F41.1 | 484 (4.8) | 6424 (4.1) | 0.0003 | 484 (4.8) | 435 (4.3) | 0.0980 |
| Wheelchair Dependence | Z99.3 | 13 (0.1) | 892 (0.6) | < 0.0001 | 13 (0.1) | 18 (0.2) | 0.3688 |
| Housing or Economic Insecurity | Z59.8 | 54 (0.5) | 740 (0.5) | 0.3375 | 53 (0.5) | 50 (0.5) | 0.7670 |
| History of PCI | 027** | 707 (7.1) | 16304 (10.4) | < 0.0001 | 707 (7.1) | 692 (6.9) | 0.6775 |
| History of CABG | 021** | 201 (2.0) | 8135 (5.2) | < 0.0001 | 201 (2.0) | 213 (2.1) | 0.5512 |
| History of Dialysis | 1012740*** | 109 (1.1) | 3612 (2.3) | < 0.0001 | 109 (1.1) | 113 (1.1) | 0.7872 |
| History of AVR | 1006141**** | 31 (0.3) | 1250 (0.8) | < 0.0001 | 31 (0.3) | 28 (0.3) | 0.6957 |

\*Includes ICD-10, ICD-10-PCS, and CPT systems as applicable
\*\*Includes all subsequent characters for available codes
\*\*\*Included codes: CPT 9093(5,7), 9094(0,5,7), 909(51–70), 90989, 9099(3,7,9).
\*\*\*\*Included codes: CPT 333(61–70), 3339(0,1), 3340(0,1), 3340(3–6), 3341(0–7), 3344
Baseline demographic and clinical characteristics are shown. Categorical data are expressed as a number (percentage), and continuous data as mean ± standard deviation. P-values indicate statistical differences between groups. After propensity score matching, both cohorts were balanced across all variables, as evidenced by nonsignificant p values ( $p > 0.05$ ).
<sup>a</sup>Includes ICD-10, ICD-10-PCS, and CPT systems as applicable
<sup>b</sup>Includes all subsequent characters for available codes
*AKI = Acute Kidney Injury; AVR = Aortic Valve Replacement; CABG = Coronary Artery*
*Bypass Graft; CKD = Chronic Kidney Disease; PCI = Percutaneous Coronary Intervention*

**Table 3.** Primary Outcomes for CTO Patients Undergoing CR.

| Outcome | aHR | 95% CI | p-value |
| --- | --- | --- | --- |
| All-Cause Mortality | 0.68 | (0.619,0.747) | < 0.0001 |
| Acute MI | 0.877 | (0.773,0.996) | 0.0429 |
| Stroke | 0.875 | (0.766,0.999) | 0.0480 |
| MACE | 0.768 | (0.698,0.846) | < 0.0001 |
| PCI | 1.005 | (0.851,1.187) | 0.9546 |
| CABG | 0.715 | (0.565,0.905) | 0.0051 |

Hazard ratios with 95% confidence intervals and p-values for various outcomes from 90 days to 5 years following CTO diagnosis are shown. CR was associated with reduced all-cause mortality, acute MI, stroke, MACE, and CABG. Difference in PCI was not statistically significant. *CABG = Coronary Artery Bypass Graft; CR = Cardiac rehabilitation; CTO = Chronic Total Occlusion; MACE = Major Adverse Cardiac Events; MI = Myocardial Infarction; PCI = Percutaneous Coronary Intervention*

**Table 4.** Baseline Characteristics of CTO Patients Starting CR Without Recent Revascularization.

|  |  | Before Propensity Score Matching |  |  | After Propensity Score Matching |  |  |
| --- | --- | --- | --- | --- | --- | --- | --- |
| Characteristics | Codes* | +CR | -CR | p-value | +CR | -CR | p-value |
| Number of Patients |  | 1,471 | 69,833 |  | 1,470 | 1,470 |  |
| Age: mean yrs ± SD (continuous) | | 68.1 ± 11.2 | 69.1 ± 11.2 | 0.0015 | 68.1 ± 11.2 | 68 ± 11.3 | 0.7514 |
| Male |  | 1089 (74.0) | 48500 (69.5) | 0.0002 | 1088 (74.0) | 1089 (74.1) | 0.9664 |
| White |  | 1167 (79.3) | 49698 (71.2) | < 0.0001 | 1166 (79.3) | 1166 (79.3) | 1.0000 |
| Black or African American |  | 132 (9.0) | 6028 (8.6) | 0.6469 | 132 (9.0) | 139 (9.5) | 0.6554 |
| Hispanic or Latino |  | 68 (4.6) | 4720 (6.8) | 0.0012 | 68 (4.6) | 57 (3.9) | 0.3147 |
| Asian |  | 37 (2.5) | 3660 (5.2) | < 0.0001 | 37 (2.5) | 39 (2.7) | 0.8162 |
| Other Race |  | 30 (2.0) | 1894 (2.7) | 0.1147 | 30 (2.0) | 33 (2.3) | 0.7024 |
| American Indian or Alaska Native |  | 10 (0.7) | 309 (0.4) | 0.1774 | 10 (0.7) | 14 (1.0) | 0.4123 |
| Native Hawaiian or Other Pacific Islander |  | 10 (0.7) | 1835 (2.6) | < 0.0001 | 10 (0.7) | 10 (0.7) | 1.0000 |
| Hypertension | I10-I15 | 1054 (71.7) | 48017 (68.8) | 0.0185 | 1053 (71.6) | 1035 (70.4) | 0.4643 |
| Hyperlipidemia | E78.5 | 840 (57.1) | 38200 (54.7) | 0.0707 | 839 (57.1) | 826 (56.2) | 0.6285 |
| Diabetes mellitus | E08-E13 | 553 (37.6) | 27192 (39.0) | 0.2912 | 552 (37.6) | 520 (35.4) | 0.2201 |
| Overweight and obesity | E66 | 379 (25.8) | 15552 (22.3) | 0.0015 | 378 (25.7) | 350 (23.8) | 0.2315 |
| Tobacco use | Z72.0 | 106 (7.2) | 4660 (6.7) | 0.4196 | 106 (7.2) | 95 (6.5) | 0.4215 |
| Obstructive sleep apnea | G47.3 | 257 (17.5) | 9795 (14.0) | 0.0002 | 257 (17.5) | 233 (15.9) | 0.235 |
| Systolic heart failure | I50.2 | 257 (17.5) | 12640 (18.1) | 0.5318 | 256 (17.4) | 243 (16.5) | 0.523 |
| Diastolic heart failure | I50.3 | 148 (10.1) | 8182 (11.7) | 0.05 | 148 (10.1) | 131 (8.9) | 0.2847 |
| Cardiomyopathy | I42 | 190 (12.9) | 9670 (13.9) | 0.3041 | 190 (12.9) | 184 (12.5) | 0.7398 |
| Atrial fibrillation and flutter | I48 | 67 (18.2) | 14715 (21.1) | 0.0064 | 267 (18.2) | 266 (18.1) | 0.9618 |
| Acute myocardial infarction | I21 | 304 (20.7) | 14241 (20.4) | 0.8009 | 303 (20.6) | 268 (18.2) | 0.1027 |
| Peripheral vascular disease | I73.9 | 180 (12.2) | 11845 (17.0) | < 0.0001 | 180 (12.3) | 179 (12.2) | 0.9551 |
| Cerebral infarction | I63 | 05 (7.1) | 5772 (8.3) | 0.119 | 105 (7.1) | 106 (7.2) | 0.943 |
| Transient cerebral ischemic attacks | G45.9 | 68 (4.6) | 2967 (4.3) | 0.4834 | 68 (4.6) | 62 (4.2) | 0.5904 |
| Chronic obstructive pulmonary disease | J44 | 188 (12.8) | 12517 (17.9) | < 0.0001 | 188 (12.8) | 181 (12.3) | 0.6968 |
| Anemia, unspecified | D64.9 | 242 (16.5) | 13286 (19.0) | 0.0125 | 242 (16.5) | 221 (15.0) | 0.2877 |
| CKD stage 3 | N18.3 | 194 (13.2) | 9616 (13.8) | 0.5189 | 194 (13.2) | 184 (12.5) | 0.5816 |
| CKD stage 4 | N18.4 | 36 (2.5) | 3293 (4.7) | 0 | 36 (2.5) | 38 (2.6) | 0.8138 |
| CKD stage 5 | N18.5 | 10 (0.7) | 1563 (2.2) | 0.0001 | 10 (0.7) | 14 (1.0) | 0.4123 |
| ESRD | N18.6 | 25 (1.7) | 3678 (5.3) | < 0.0001 | 25 (1.7) | 24 (1.6) | 0.8855 |
| Osteoarthritis | M15-M19 | 395 (26.9) | 16570 | 0.0055 | 395 (26.9) | 389 (26.5) | 0.8024 |
|  |  |  | (23.7) |  |  |  |  |
| Dementia | F03 | 14 (1.0) | 1417 (2.0) | 0.0035 | 14 (1.0) | 14 (1.0) | 1.0000 |
| Depression | F32 | 237 (16.1) | 9881 (14.2) | 0.0332 | 236 (16.1) | 236 (16.1) | 1.0000 |
| Generalized anxiety disorder | F41.1 | 58 (3.9) | 2700 (3.9) | 0.8819 | 58 (4.0) | 51 (3.5) | 0.4944 |
| Wheelchair Dependence | Z99.3 | 10 (0.7) | 459 (0.7) | 0.9164 | 10 (0.7) | 10 (0.7) | 1.0000 |
| Housing or Economic Insecurity | Z59.8 | 12 (0.8) | 325 (0.5) | 0.0526 | 11 (0.8) | 10 (0.7) | 0.8266 |
| History of PCI | 027** | 10 (0.7) | 526 (0.8) | 0.7463 | 10 (0.7) | 10 (0.7) | 1.0000 |
| History of CABG | 021** | 0 (0.0) | 10 (0.0) | 0.6462 | 0 (0.0) | 0 (0.0) | -- |
| History of Dialysis | 1012740*** | 11 (0.8) | 1400 (2.0) | 0.0006 | 11 (0.8) | 13 (0.9) | 0.6819 |
| History of AVR | 1006141**** | 10 (0.7) | 300 (0.4) | 0.1491 | 10 (0.7) | 10 (0.7) | 1.0000 |

\*Includes ICD-10, ICD-10-PCS, and CPT systems as applicable
\*\*Includes all subsequent characters for available codes
\*\*\*Included codes: CPT 9093(5,7), 9094(0,5,7), 909(51–70), 90989, 9099(3,7,9).
\*\*\*\*Included codes: CPT 333(61–70), 3339(0,1), 3340(0,1), 3340(3–6), 3341(0–7), 3344
Baseline demographic and clinical characteristics are shown. Categorical data are expressed as a number (percentage), and continuous data as mean ± standard deviation. P-values indicate statistical differences between groups. After propensity score matching, both cohorts were balanced across all variables, as evidenced by nonsignificant p values ( $p > 0.05$ ).
<sup>a</sup>Includes ICD-10, ICD-10-PCS, and CPT systems as applicable
<sup>b</sup>Includes all subsequent characters for available codes
*AKI = Acute Kidney Injury; AVR = Aortic Valve Replacement; CABG = Coronary Artery*
*Bypass Graft; CKD = Chronic Kidney Disease; PCI = Percutaneous Coronary Intervention*

**Table 5.** Primary Outcomes for CTO Patients Undergoing CR Without Recent Revascularization.

| Outcome | aHR | 95% CI | p-value |
| --- | --- | --- | --- |
| All-Cause Mortality | 0.814 | (0.664,0.996) | 0.0455 |
| Acute MI | 1.049 | (0.787,1.397) | 0.7460 |
| Stroke | 1.142 | (0.824,1.583) | 0.4238 |
| MACE | 0.905 | (0.731,1.12) | 0.3574 |
| PCI | 1.315 | (0.98,1.763) | 0.0669 |
| CABG | 0.439 | (0.245,0.785) | 0.0043 |

Hazard ratios with 95% confidence intervals and p-values for various outcomes from 90 days to 5 years following CTO diagnosis without prior revascularization are shown. CR was associated with reduced all-cause mortality. No differences were found in MI, stroke, CABG, PCI, or MACE. *CABG = Coronary Artery Bypass Graft; CR = Cardiac rehabilitation; CTO = Chronic Total Occlusion; MACE = Major Adverse Cardiac Events; MI = Myocardial Infarction; PCI = Percutaneous Coronary Intervention*

## Results

Of 167,176 CTO patients, 10,021 enrolled in CR, including 1,608 patients without another CR indication before matching. After matching, there were 10,020 patients in each cohort for the primary analysis, with 1,608 patients in each cohort for the secondary analysis. Baseline characteristics for the primary analysis before and after propensity matching for 39 unique variables demonstrated adequate balancing, evidenced by p-values <0.05 for all but 3 characteristics and standardized mean differences <0.05 for all characteristics (Table 2).

In the primary analysis, participation in CR was significantly associated with reduced all-cause mortality (HR 0.68; 95% CI, 0.62–0.75; p < 0.0001), acute MI (HR 0.87; 95% CI, 0.77–1.00; p = 0.043), stroke (HR 0.88; 95% CI, 0.77–1.00; p = 0.048), MACE (HR 0.77; 95% CI, 0.70–0.85; p < 0.0001), and revascularization with CABG (HR 0.72; 95% CI, 0.57–0.91; p = 0.005) at 5 years. No significant differences were found for PCI in the primary analysis.

In the secondary analysis, which excluded patients with prior revascularization, CR participation remained associated with significantly reduced all-cause mortality (HR 0.81; 95% CI, 0.67–0.99; p < 0.036) at 5 years. However, no significant differences were observed for acute MI, stroke, MACE, need for CABG, and PCIs.

## Discussion

This large, propensity-matched cohort study suggests an association between cardiac rehabilitation (CR) and a significant reduction in 5-year all-cause mortality among patients with coronary chronic total occlusions (CTOs). We observed a 26.7% relative risk reduction for CR participants vs. non-CR patients (absolute risk 7.7% vs. 10.5%, aHR 0.68). In patients managed with OMT alone, CR was associated with a 10.4% relative risk reduction compared to no CR (absolute risk 12.4% vs. 13.9%, aHR 0.81). These findings align with Class 1 secondary prevention recommendations for CR but extend them specifically to patients with CTOs, a population poorly represented in prior trials. ^10^.

The magnitude of benefit with CR observed in this study exceeds what is typically reported in large CR RCTs for broad coronary artery disease populations. In an updated Cochrane meta-analysis of 85 CR RCTs, no effect on all cause mortality was found despite a 26% reduction in cardiovascular mortality ^14^. Among the CTO population specifically, contemporary RCTs—such as EuroCTO and DECISION-CTO—have also failed to demonstrate a significant impact of CTO PCI on major adverse cardiac events or mortality, despite improvements in angina relief and quality of life^17^,^18^. Our study found a 26.7% relative risk reduction in all-cause mortality for all new CTO patients and a 10.4% relative risk reduction for patients managed via MT alone. As the presence of CTOs are associated with higher mortality there is strong need for therapies that can provide mortality benefit in this population^2^,^15^,^16^. Our findings highlight how CR may provide an \noninvasive, \benefit in a high-risk patient group. This may position CR as a valuable adjunct to optimal medical therapy, particularly for patients in whom complete revascularization is not feasible or expected to confer survival benefit.

The potential mechanisms for this observed association are likely multifactorial. Participation in CR has been shown to improve systemic endothelial function, restore sympathovagal balance, and exert anti-inflammatory effects^22^⁻^25^. Furthermore, CR is a multidisciplinary intervention that provides a structured platform for reinforcing adherence to OMT and delivering psychosocial support^26^.

The primary strength of this study includes the use of large, real-world, propensity-matched cohorts that minimize confounding to provide statistical power. Our landmark analysis further mitigated immortal time bias by aligning follow-up between exposure groups. Importantly, CR is scalable, noninvasive, and already reimbursed, making it a pragmatic strategy for immediate implementation while definitive trials are pursued. These findings underscore the need for a large-scale, adequately powered randomized controlled trial with extended follow-up to definitively assess the long-term clinical impact of CR in this complex and high-risk population.

## Limitations

The main limitation of our study remains its retrospective and observational nature. Although our propensity matching was considered balanced, we cannot eliminate the risk of unmeasured confounding and selection bias. Patients who participate in CR may represent a more motivated cohort with healthier baseline lifestyle behaviors, stronger social support, or lower frailty not fully captured in our dataset. Additionally, CR participation was defined based on billing code documentation, which cannot fully capture adherence, quality, or duration of CR programs. Finally, we were unable to account for angiographic complexity, symptom burden, and medication adherence in our dataset, which may limit the generalizability of our results.

## Conclusion

This large-scale, real-world analysis demonstrates a strong association between CR participation and reduced 5-year mortality in patients with CTOs. The magnitude of this association underscores the potential of CR as a powerful, low-risk adjunct to optimal medical therapy in a population where invasive revascularization strategies carry substantial procedural risks with limited evidence on long-term outcome benefits. These findings support the need for adequately powered, prospective randomized trials to definitively evaluate the impact of CR in this complex, high-risk patient population and clarify its role in their long-term management.

## Data Availability

The data that support the findings of this study are available from TriNetX but restrictions apply to the availability of these data, which were used under license for the current study, and so are not publicly available. Data are available from the authors upon reasonable request and with permission of TriNetX

## Non-standard Abbreviations and Acronyms

CABG: Coronary Artery Bypass Graft
CAD: Coronary Artery Disease
CR: Cardiac Rehabilitation
CTO: Chronic Total Occlusions
MACE: Major Adverse Cardiac Events
MI: Myocardial Infarction
PCI: Percutatenous Coronary Intervention
ICD: International Classification of Disease, Tenth Edition

## Acknowledgements

Artificial intelligence software (ChatGPT, OpenAI) was used to assist with drafting and editing portions of the manuscript text and in compiling sections of the report. No AI tools were used for the study design, data analysis, or performance of the research

